# Effects of plasmapheresis frequency on health status and exercise performance in men: a randomized controlled trial

**DOI:** 10.1101/2023.06.05.23290943

**Authors:** Alexandre Mortier, Jina Khoudary, Sophie van Doorslaer de Ten Ryen, Camille Lannoy, Nicolas Benoit, Nancy Antoine, Sylvie Copine, Hans Van Remoortel, Philippe Vandekerckhove, Veerle Compernolle, Louise Deldicque

## Abstract

**Objectives:** To collect data on 1) haematological and biochemical markers, 2) physiological and exercise-related parameters and 3) adverse events to get a comprehensive picture of the safety of intensive or less intensive plasma donation protocols.

**Methods:** Sixty-three male subjects participated in this randomized controlled trial and were divided into a low-frequency (LF, 1x/month, n=16), high-frequency (HF, 3x/month, n=16), very high-frequency (VHF, 2x/week, n=16) and a placebo (P, 1x/month, n=15) group. Biochemical, haematological, clinical, physiological and sport performance-related data were collected before (D0), after 1½ month (D42) and after 3 months (D84).

**Results:** In VHF, red blood cells, haemoglobin and haematocrit levels decreased while reticulocyte levels increased from D0 to D84. In both HF and VHF, plasma ferritin levels were lower at D42 and D84 compared to D0. In VHF, the plasma levels of albumin, IgG, IgA and IgM dropped from D0 to D42 and remained lower at D84 than at D0. In HF, plasma IgG, IgA and IgM were lower at D42 and IgG and IgM were lower at D84 compared to D0. Few adverse events (haematoma, anaemia, vasovagal reactions without syncope) were reported in HF and VHF. Repeated plasma donation had no effect on blood pressure, body composition or exercise performance.

**Conclusion:** Haematological and biochemical parameters were severely impacted when plasmapheresis was repeated twice a week, mildly impacted with a frequency of three times per month and not impacted with a frequency of once a month. Despite those changes, markers for exercise performance were not altered when plasmapheresis, whatever the frequency, was repeated over 3 months.

**Summary Box:** *What is already known on this topic?:* Plasmapheresis may induce a series of health consequences, such as adverse events of diverse severities. However, those events have mainly been studied separately and retrospectively with no appropriate placebo group.

*What this study adds?:* This is the first randomized controlled trial prospectively investigating the effects of repeated plasma donation on a whole range of health consequences, namely biochemical and haematological, blood pressure, body composition, adverse events and, exercise performance.

*How this study might affect research, practice or policy?:* Worldwide efforts need to be made to increase the amount of plasma collected for manufacturing plasma-derived product. To avoid negative effects to donors, a sustainable plasma supply should be based on a large donorbase of low-frequency donors.

## INTRODUCTION

More and more inflammatory, neurological, haematological, and immunological diseases can be effectively treated with human plasma-derived products.^1^ As such, the demand for plasma as the starting material for the manufacture of intravenous immunoglobulin and other plasma derivatives is growing significantly and is expected to continue to increase.^2^ Future demand for intravenous immunoglobulin in developed countries is largely being driven by populations that are increasing in age and weight as well as the emergence of new indications. To increase the amount of collected plasma, one can expand the existing donorbase, collect higher volumes per donation or stimulate donors to donate more frequently. Increasing plasma collections, whether by increasing the volume per donation or the frequency, from returned donors can also reduce the risk of infectious disease transmission, as these donors have been shown to have a lower incidence of infectious disease markers.^3 4^

Plasma donors in the United States may donate twice within 7 days as long as the interval between donations is at least 2 days.^5^ Prospective studies conducted in Germany, such as the safety of long-term intensive plasmapheresis (SIPLA) study, have demonstrated that up to 850ml of plasma, including anticoagulant, can be safely collected.^6^ However, only very few studies have prospectively looked at the effect of repeated intensive plasma donation and whether a control group was lacking^6 7^ or the control group was not randomized.^8^ The majority of the studies were retrospective^9–14^ or limited to one single donation^15–17^. We have recently found that repeated whole blood donation with a 3-month interval in between induced a drop in markers for iron status, which worsened with the number of donations.^18^ The repetition effect of the donations, whether whole blood or plasma, can be different from the effects measured after one single donation. It is therefore critical to test and document this repetitive effect to build trustable and valid guidelines concerning repetitive plasma donation. Up to now, each study has looked at a quite limited number of outcomes separately, namely biochemical and haematological,^6 8 10–13 15 16^ blood pressure,^7^ clinical symptoms and adverse events,^14 15^ bone metabolism,^9^ and exercise performance.^17^ The aim of the present study was to collect data on 1) haematological and biochemical markers, 2) physiological and exercise-related parameters and 3) adverse events to get a comprehensive picture of the safety of intensive or less intensive plasma donation protocols over a 3-month period.

## MATERIAL AND METHODS

### Equity, diversity, and inclusion statement

The author group is gender balanced and consists of junior, mid-career and senior researchers from different disciplines; however, all members of the author group are from one country. Our study population included adult men from different socioeconomic backgrounds; thus, findings may not be generalizable to other populations. The influence of gender and age is considered in the discussion.

### Subjects

Potential study participants were either new plasmapheresis donors or previous donors who had not donated for at least 2 weeks. Eligible study participants were randomly assigned, via a computer-generated randomization table, into a placebo group (P), a low-frequency group (LF, 1 plasma donation per month), a high-frequency frequency group (HF, 3 plasma donations per month) and a very high-frequency group (VHF, 2 plasma donations per week), to participate in this longitudinal study. The donation frequency regimen in the VHF group (2x/week) represents the current plasma donation regulation in countries such as Austria, Germany, Hungary, USA, and Canada whereas most other countries have a minimal donation interval of 14 days.^19^ The placebo group underwent a placebo donation at the same frequency as LF (1x/month). All participants were blinded to the group selection.

Inclusion criteria were as follows: male, age 18 to 50 y, body mass index (BMI) 20 to 28 kg×m^2-1^, and no contraindication to perform maximal intensity exercise assessed by the physical activity readiness questionnaire (PAR-Q). During the whole duration of the study, subjects were asked to maintain their habitual lifestyle, i.e., physical activity and diet. All participants provided written informed consent after the explication of all potential risks of the study and the right to withdraw at any time. This study was conducted at the UCLouvain in Belgium from March 2022 to December 2022. The study was approved by the Ethics Committee of UCLouvain and the investigation was performed according to the principles outlined in the Declaration of Helsinki. The study (RK2020) was registered at clinicaltrials.gov and received the identifier NCT05815615.

### Experimental procedures

One week before the first plasma donation (D0, visit 1), subjects reported to the exercise physiology laboratory. First, systolic and diastolic blood pressure was measured automatically (Omron) in the supine position. Then, five blood samples were taken in an antecubital vein from the non-dominant arm: three 4-ml EDTA tubes, one 8-ml clot activator tube and one 4-ml sodium fluoride/potassium oxalate tube. After blood sampling, a maximal strength test of the dominant arm was performed using an electronic dynamometer (Grip-D, Takei, Japan) followed by a maximal strength test (1RM) with the dominant leg on a leg extension machine (ProDual, Body-Solid, IL, USA). After an individualised and standardised warm-up protocol, the 1RM test consisted of a maximum of 5 attempts interspersed by 3-min rest. The 1RM corresponded to the highest load lifted once with a correct technique. Then, body mass and height were measured (Seca GmbH., Hamburg, Germany) and body composition (bone mineral content, fat-free mass and fat mass) was assessed by dual energy X-ray absorptiometry (DXA) scan (Discovery W, Hologic Inc., MA, USA). Finally, a progressive test on a bicycle ergometer (Cyclus 2, RBM elektronik automation GmbH, Leipzig, Germany) was performed to measure peak oxygen consumption (VO_2_ peak). The test started at 70 watts (W), followed by incremental loads of 30 W every 2 min until exhaustion. The maximal power output (Pmax) was calculated as the last step completed plus the last increment corrected for the sustained duration, which corresponded to the total time of the test. VO_2_, carbonic dioxide production (VCO_2_) (Medisoft Ergocard, MGC Diagnostics Corporation, MN, USA) as well as heart rate (HR) (Polar, Kempele, Finland) were continuously monitored during the test. Oxygen pulse was calculated by dividing VO_2_ peak and HRmax at the end of the test. Blood lactate was measured before, during (at 190 W), and at the end of the test by taking a capillary blood sample (5 μl) from an earlobe (Lactate Pro, Arkray, Japan). The whole sampling and testing procedure was repeated 42 days (D42, visit 2) and 84 days (D84, visit 3) after the first plasma donation in exactly the same conditions. Adverse events, categorized according to international standards^20^, were recorded in the blood information system throughout the whole experimental trial. Citrate reactions were not considered in our analysis, since preventive calcium supplements were provided standardly to the regular donors.

### Plasma donation

One week after blood sampling and exercise pretesting (D0 or visit 1), participants reported to the Red Cross Center in Leuven or Mechelen (Belgium). They underwent a plasma donation of 650 ml (exclusive anticoagulant) according to the Belgian Law of 01/02/2005, without exceeding 20% of total body volume during or 16% of total body volume at the end of the plasma donation (donation group), or had a similar sensation of undergoing a plasma donation (placebo group), by infusing NaCl 0.9% using a NexSys PCS device (Haemonetics). During each donation or simulation of donation, the puncture arm was shielded, and subjects were listening to music through a headset.

### Blood analyses

Each tube was centrifuged for 10 min at 2000 g at 4°C and analyzed within 24 h following blood drawing. The supernatant was collected and stored at -80 °C. The following parameters were analyzed in a medical analysis laboratory (Lims mbnext group Europe, LLN, Belgium): red blood cell, haemoglobin, haematocrit, reticulocyte, iron, ferritin, C-reactive protein (CRP), glycemia, insulinemia, glycated haemoglobin (HbA1c), creatine kinase (CK), total cholesterol, albumin, immunoglobulin (Ig) A, IgG and IgM.

### Statistical analysis

A statistical power analysis was performed to determine the optimal number of subjects needed to find a difference in total serum protein mean of 10%^10^ with a standard deviation corresponding to 8% and a power of 80% according to the calculator developed by Wang and Ji. ^21^ Drop-outs were not taken into consideration in statistical analysis. Potential differences in subjects’ characteristics at baseline were analyzed with a one-way ANOVA (IBM SPSS Statistics, Version 28.0., Armonk, NY, USA). A mixed ANOVA model for repeated measures (SAS Statistical Software 9.4, SAS Institute, Cary, NC, USA) was used with the subjects as a random variable and groups (placebo and donations) and condition (time) as fixed independent variables. The p-values of the main effects can be found in Table S1. The model used Kenward-Roger approximation of degree of freedom with compound symmetry variance-covariance structure. When appropriate, contrast analyses were performed to compare means, applying a Sidak correction. Linear mixed models for repeated measures give unbiased results in the presence of missing data and take potential differences at baseline into account. Normality of residuals was tested using a QQ plot. The total amount of adverse events and the adverse event rate (per 50 donations) were calculated. Statistical significance was set at p<0.05. All values are expressed as mean ± SEM, except donation history as a median [IQR, interquartile range].

## RESULTS

### Subject characteristics

Seventy-two volunteers were enrolled in the study: 17 in the Placebo, 16 in the LF, 20 in the HF and 19 in the VHF group (Fig. 1). Nine subjects (2 in the placebo group, 4 in the HF group and 3 in the VHF group) gave up before the end of the study. Withdrawal from the study was due to personal reasons or impossibility to comply with the repeated appointments. As they were incomplete, the data of the drop-outs were not included in the analyses, resulting in a total number of 63 subjects: 15 in the Placebo, 16 in the LF, 16 in the HF and 16 in the VHF group. All participants were regular donors, except one new donor (who was allocated to the LF group). At the start of the study, the 4 groups were not different regarding age, BMI, the amount of physical activity per week, VO_2_peak and donation history (Table S2).

**Fig. 1.**
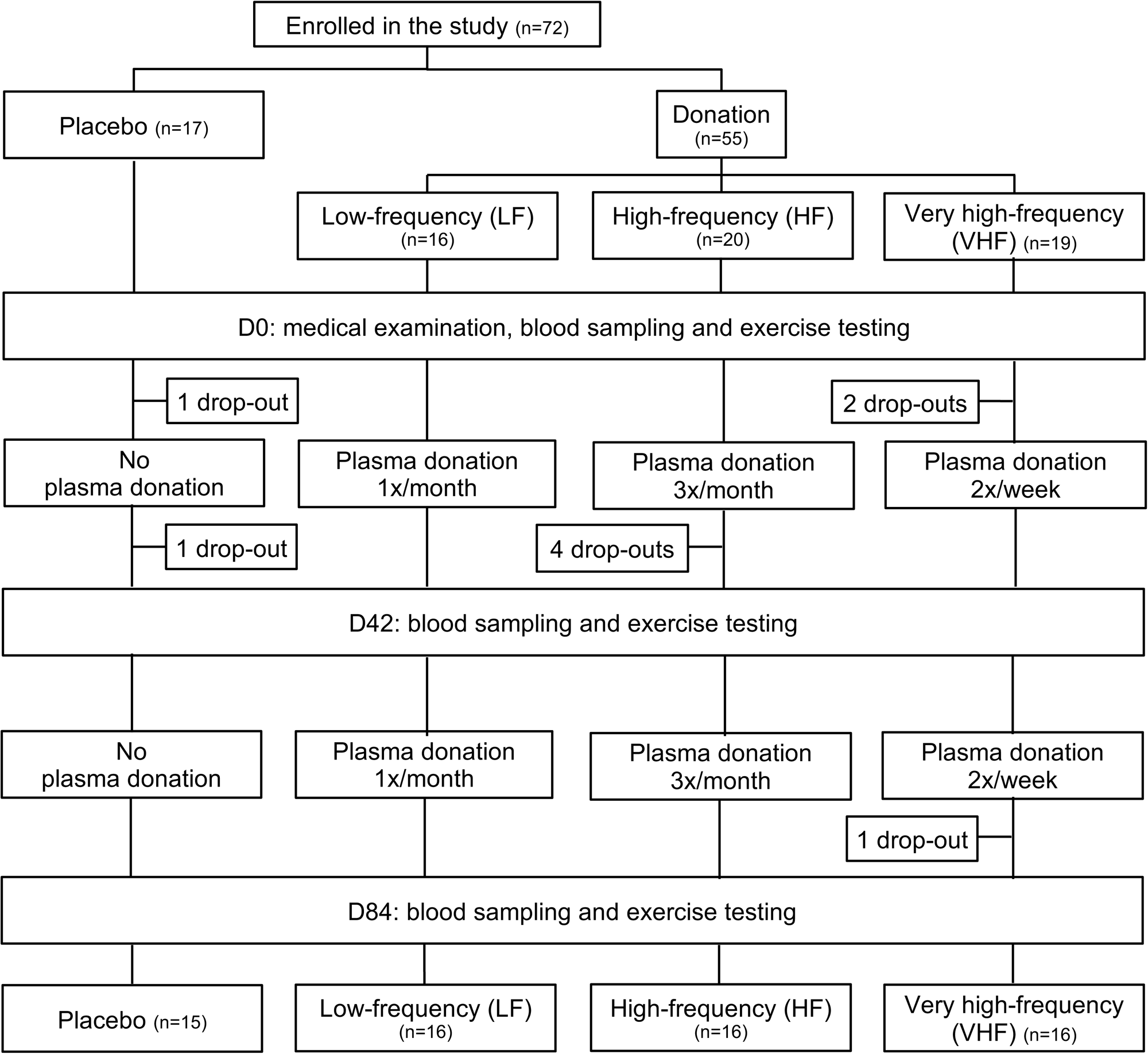
Subjects flow chart.

Forty-nine study participants were 100% compliant to the corresponding donor regimen (5 (31%) in the VHF group, 14 (88%) in the HF group, 15 (94%) in the LF group, and 15 (100%) in the placebo group). In het VHF group, 6 participants missed 1 donation, 2 participants missed 2-3 donations, 2 participants missed 5-6 donations, and 1 participant missed 10 donations.

### High-frequency and very high-frequency donation affect haematology

Red blood cells, haemoglobin, haematocrit, reticulocyte, iron and ferritin levels all evolved differently between the 4 groups after repeated plasma donation (main group or main group*time effect, p<0.05-0.001, Table S1 and Figure 2A and B). In VHF, red blood cells (p<0.001), haemoglobin (p<0.001) and haematocrit (p=0.003) levels decreased while reticulocytes levels (p<0.001) increased from D0 to D84 (Table 1 and Figure 2A and B). In addition, reticulocytes levels were higher at D42 compared to D0 (p<0.001). In both HF and VHF, plasma ferritin levels were lower at D42 (p=0.039 in HF; p=0.001 in VHF) and at D84 (p=0.008 in HF; p<0.001 in VHF) compared to D0. Except for red blood cells, all aforementioned effects of repeated plasma donation in HF and VHF were different from P at the same time point.

**Fig. 2.**
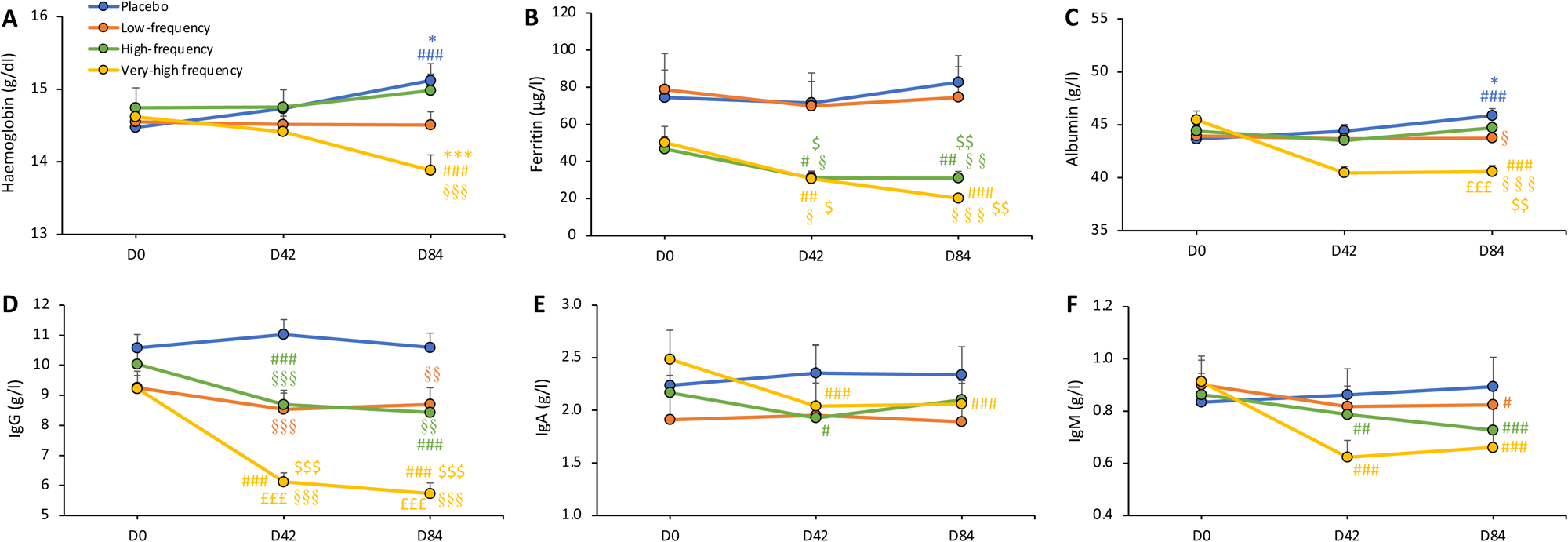
Plasma haemoglobin, ferritin, albumin and immunoglobulin levels. Evolution of plasma haemoglobin (A), ferritin (B), albumin (C), Immunoglobulin G (IgG, D), IgA (E) and IgM (F) levels before (D0), during (D42) and after (D84) the 3-month trial in the placebo and donation groups. Data are expressed as means ± SEM. n=15 in Placebo, n=16 in each of the 3 donation groups. ^#^p<0.05, ^##^p<0.01, ^###^p<0.001 different from D0, same group. *p<0.05, ***p<0.001 D84 vs D42, same group. ^§^p<0.05, ^§§^p<0.01, ^§§§^p<0.001 different from P, same time. ^$^p<0.05, ^$$^p<0.01, ^$$$^p<0.001 different from LF, same time. ^£££^p<0.001 different from HF, same time.

**Table 1.**
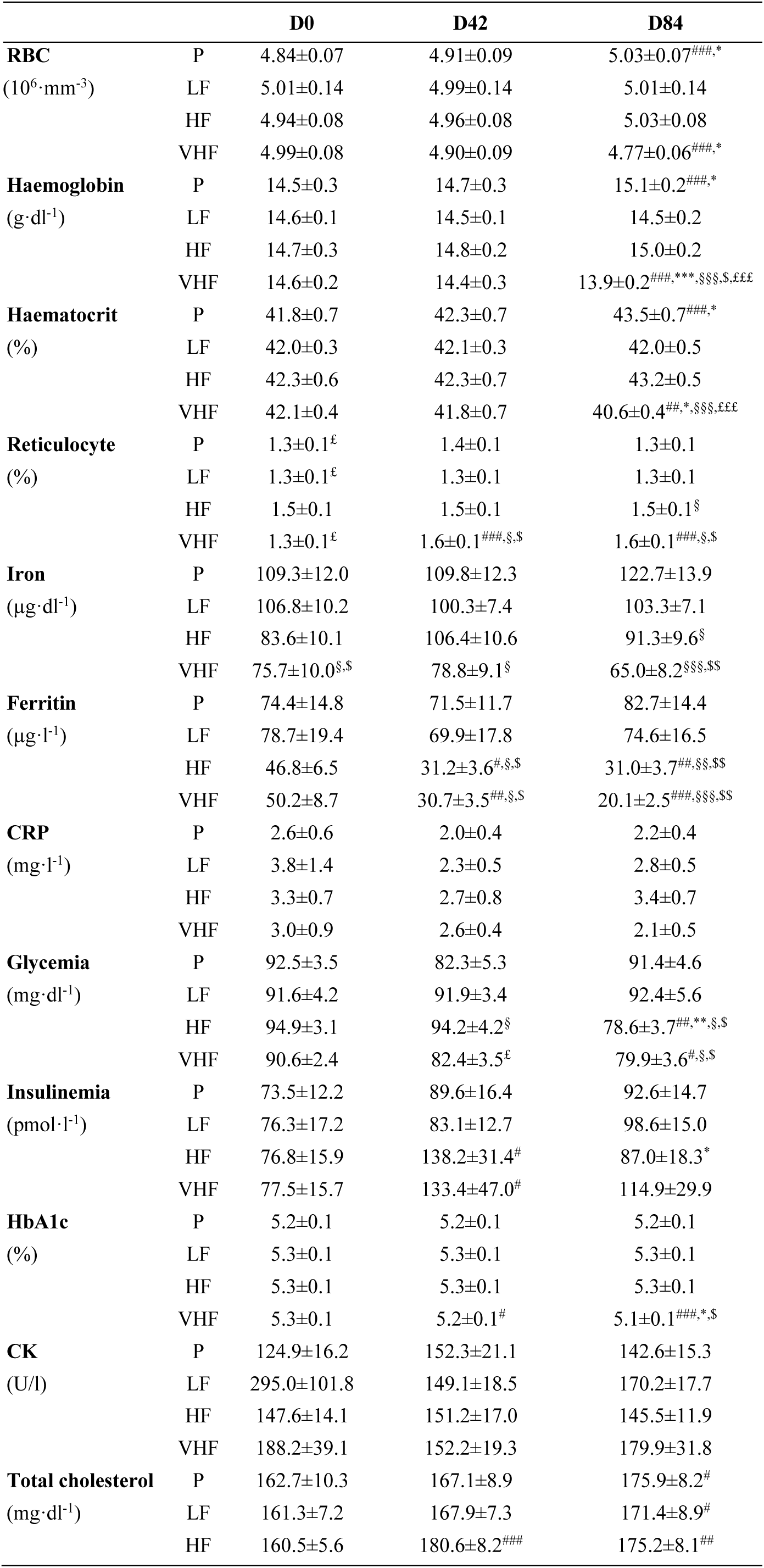
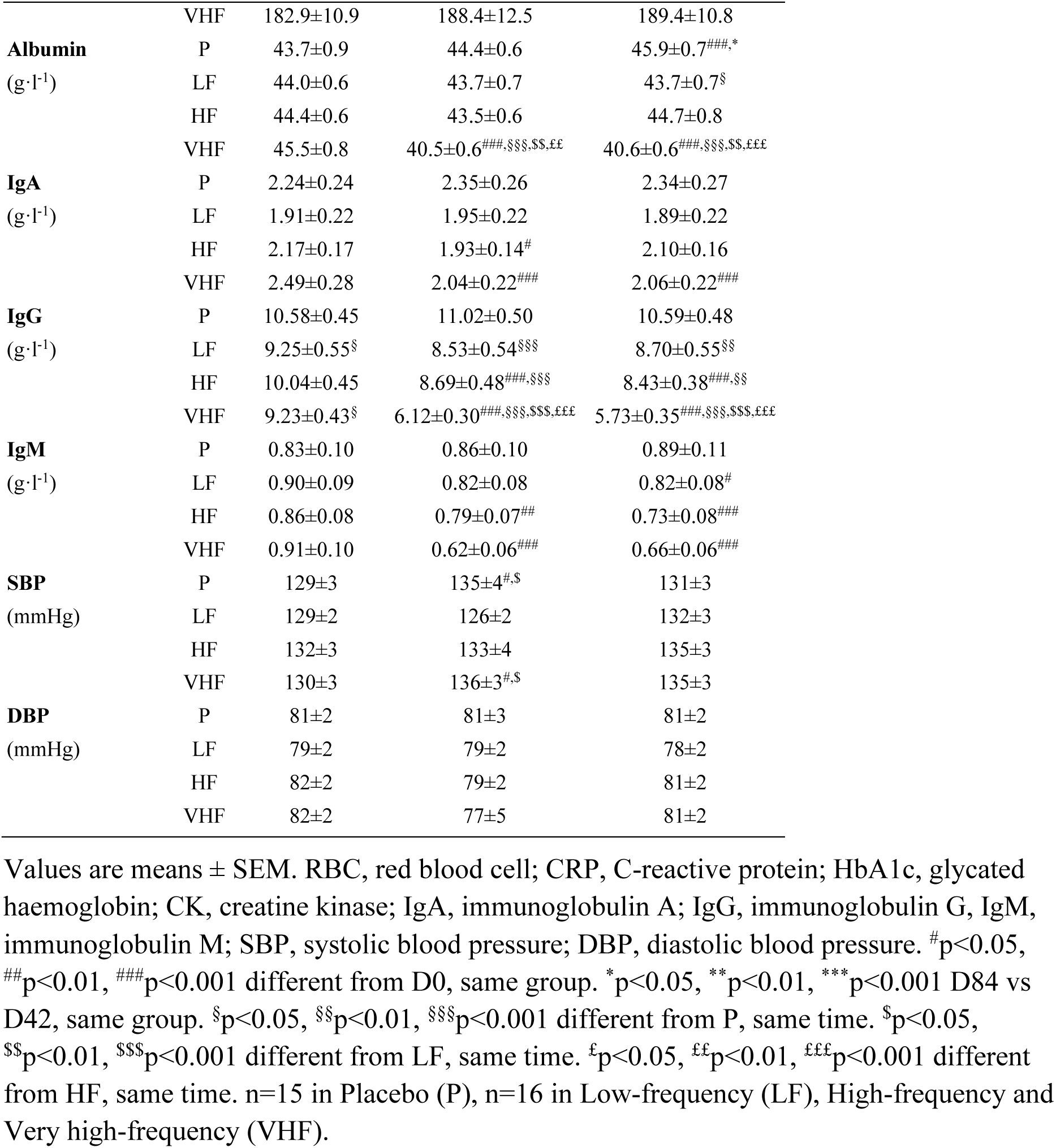
Effects of repeated plasma donation on hematological parameters and blood pressure.

### Very high-frequency donation lowers HbA1c

A main time and/or a group*time effect was present for glycemia, insulinemia and HbA1c (p<0.05-0.001, Table S1). Glycemia decreased from D0 to D84 in HF (p=0.002) and in VHF (p=0.042) and was lower in both HF and VHF at D84 compared to P and LF (p<0.05) (Table 1). Insulinemia increased from D0 to D42 in HF (p=0.035) and VHF (p=0.019). In VHF, plasma HbA1c levels decreased from D0 to D42 (p=0.007) and from D42 to D84 (p=0.015). A main time effect was found for plasma total cholesterol levels (p<0.001). Compared to D0, total cholesterol levels were higher at D42 in HF (p<0.001) and at D84 in P (p=0.014), LF (p=0.049) and HF (p=0.005).

### Very high-frequency scheme severely affects plasma albumin and immunoglobulin levels

Plasma albumin, IgG, IgA and IgM levels all evolved differently between the 4 groups after repeated plasma donation (main group*time effect, p<0.001, Table S1 and Figure 2C-F). In VHF, the plasma levels of albumin, IgG, IgA and IgM dropped substantially from D0 to D42 (p<0.001) and remained lower at D84 than at D0 (p<0.001) (Table 1). Albumin and IgG levels at D42 and D84 in VHF were lower than P, LF and HF at the same time (p<0.01-0.001). In HF, compared to D0, plasma IgG (p<0.001), IgA (p=0.011) and IgM (p=0.006) were lower at D42 and IgG (p<0.001) and IgM (p<0.001) lower at D84. CRP, CK, SBP and DBP were unaffected by repeated plasma donation.

### No effect of repeated plasma donation on body composition

A main time, but no group nor group*time, effect was found for fat free mass (p=0.012), fat free mass + bone mineral content (p=0.012), fat mass (p<0.001) and % fat (p<0.001) (Table S1). Fat free mass and fat free mass + bone mineral content was lower at D84 compared to D0 (p=0.040 and p=0.049, respectively) and D42 (p=0.008 and p=0.009, respectively) in P (Table 2). Fat mass increased from D0 to D84 in HF (p=0.038) and VHF (p=0.041). In addition, fat mass increased from D42 to D84 in P (p=0.016) and VHF (p=0.002), which resulted in an increase in fat percentage between D42 and D84 in P (p=0.006) and VHF (p=0.003). No main effect was found for body mass, BMI and bone mineral content.

**Table 2.** Effects of repeated plasma donation on body composition.

|  |  | D0 | D42 | D84 |
| --- | --- | --- | --- | --- |
| **Body mass**<br>(kg) | P | 77.4±3.8 | 77.7±3.8 | 77.2±3.7 |
|  | LF | 78.9±2.6 | 79.1±2.7 | 79.5±2.5 |
|  | HF | 82.8±2.1 | 83.7±2.5 | 83.5±2.5 |
|  | VHF | 80.7±2.9 | 80.2±2.8 | 80.7±3.0 |
| **BMI**<br>(kg·m<sup>-2</sup>) | P | 23.6±0.9 | 23.7±0.9 | 23.6±0.9 |
|  | LF | 23.7±0.6 | 23.6±0.6 | 23.8±0.7 |
|  | HF | 24.4±0.7 | 24.7±0.8 | 24.7±0.8 |
|  | VHF | 24.0±0.6 | 23.8±0.5 | 24.0±0.6 |
| **BMC**<br>(kg) | P | 2.87±0.12 | 2.88±0.12 | 2.91±0.12 |
|  | LF | 2.97±0.13 | 2.95±0.13 | 2.98±0.13 |
|  | HF | 2.95±0.06 | 2.95±0.05 | 2.93±0.06** |
|  | VHF | 2.89±0.11 | 2.87±0.11 | 2.88±0.11 |
| **Fat mass**<br>(kg) | P | 12.7±1.9 | 12.4±1.8 | 13.3±1.9* |
|  | LF | 11.5±1.1 | 11.5±1.1 | 11.8±1.1 |
|  | HF | 14.6±1.4 | 15.1±1.5 | 15.4±1.5<sup>#</sup> |
|  | VHF | 14.1±1.3 | 13.7±1.3 | 14.9±1.4<sup>#,**</sup> |
| **FFM**<br>(kg) | P | 60.6±2.3 | 60.9±2.2 | 59.8±2.1<sup>#,**</sup> |
|  | LF | 63.2±1.7 | 63.2±1.8 | 63.2±1.7 |
|  | HF | 63.9±1.2 | 64.2±1.2 | 63.6±1.2 |
|  | VHF | 61.8±2.0 | 61.9±1.8 | 61.5±1.9 |
| **FFM + BMC**<br>(kg) | P | 63.5±2.4 | 63.8±2.3 | 62.7±2.2<sup>#,**</sup> |
|  | LF | 66.1±1.8 | 66.1±1.9 | 66.2±1.7 |
|  | HF | 66.8±1.2 | 67.2±1.2 | 66.6±1.2 |
|  | VHF | 64.7±2.1 | 64.8±1.9 | 64.3±1.9 |
| **% Fat** | P | 15.9±1.6 | 15.5±1.6 | 16.8±1.6** |
|  | LF | 14.6±1.0 | 14.5±1.0 | 14.9±1.0 |
|  | HF | 17.5±1.3 | 17.9±1.4 | 18.3±1.4 |
|  | VHF | 17.7±1.2 | 17.1±1.2 | 18.4±1.2** |

Values are means ± SEM. BMI, body mass index; BMC, bone mineral content; FFM, fat free mass. <sup>#</sup>p<0.05 vs D0, same group. \*\*p<0.01 vs D42, same group. n=15 in Placebo (P), n=16 in Low-frequency (LF), High-frequency (HF) and Very high-frequency (VHF).

### No effect of repeated plasma donation on exercise performance

A main time, but no group nor group*time, effect was found for maximal power output (p=0.006, Fig. 3A), VO_2_peak (p=0.001, Fig. 3B) and oxygen pulse (p=0.002) (Table S1). Maximal power output decreased from D0 to D42 in LF (p=0.019) and from D0 to D42 (p=0.013) and D84 (p=0.012) in VHF (Fig. 3A and Table 3). VO_2_peak was higher at D42 compared to D0 (p=0.009) and compared to D84 (p=0.038) in HF (Fig. 3B). Finally, oxygen pulse was higher at D42 in P (p=0.037) and HF (p=0.006) and higher at D84 (p=0.027) in LF compared to D0. No main effect was found for lactate at 190W, lactate post-exercise, maximal ventilation and maximal heart rate, maximal quadriceps (Fig. 3C) and arm strength.

**Fig. 3.**
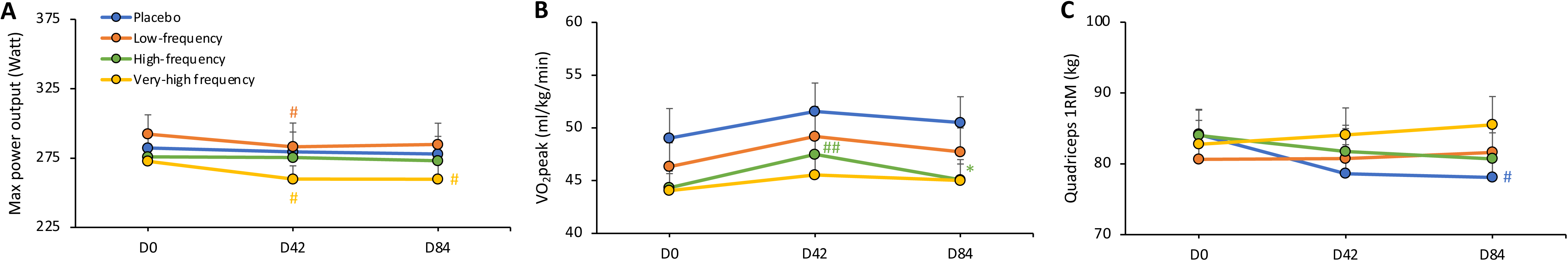
Markers for endurance and strength performance. Evolution of the maximal power output (A), peak oxygen consumption (VO_2_peak, B) and maximal strength of the quadriceps (1RM, C) before (D0), during (D42) and after (D84) the 3-month trial in the placebo and donation groups. Data are expressed as means ± SEM. n=15 in Placebo, n=16 in each of the 3 donation groups. ^#^p<0.05, ^##^p<0.01 different from D0, same group. *p<0.05 D84 vs D42, same group.

**Table 3.**
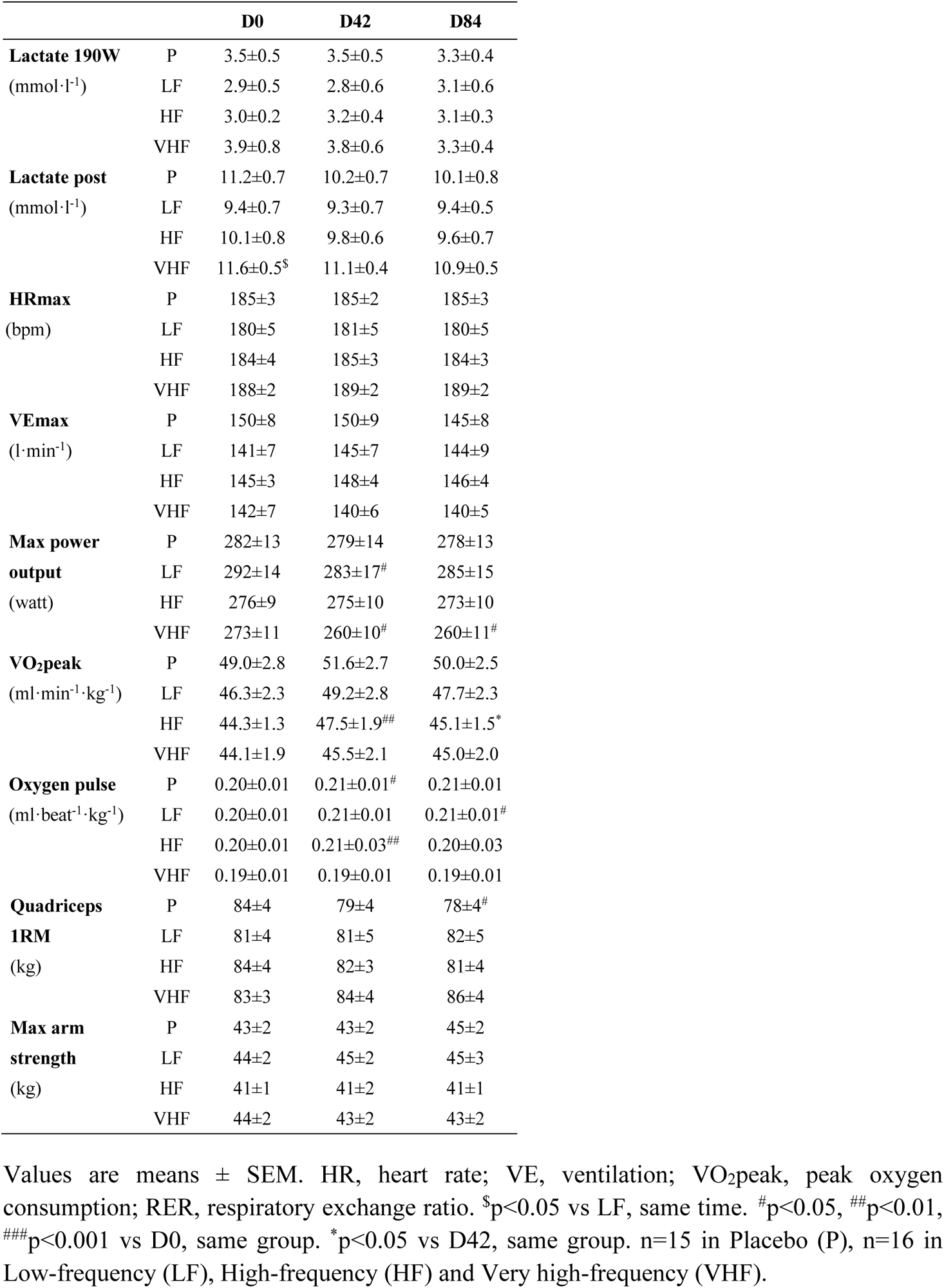
Effects of repeated plasma donation on exercise performance.

### Few adverse events were reported in the HF and VHF group

The occurrence of adverse events was monitored in each group during the whole experimental trial (Table S3). Five haematomas were present in HF (3 events in 3 donors, adverse event rate: 1.08) and VHF (2 events in 1 donor, adverse event rate: 0.28). A total amount of 5 vasovagal reactions were reported, 1 in HF (1 donor, adverse event rate: 0.36) and 4 in VHF (3 donors, adverse event rate: 0.57). Six anaemia events (in 5 donors, adverse event rate: 0.85) were detected in VHF. No other (major) events were reported.

## DISCUSSION

For the first time, data on 1) haematological and biochemical markers, 2) physiological and exercise-related parameters and 3) adverse events were prospectively collected over 3 months to get a comprehensive picture of the health consequences of intensive or less intensive plasma donation protocols. We found that repeated plasma donation: 1) decreased red blood cells, haemoglobin and haematocrit levels and increased reticulocytes levels in the very high-frequency group; 2) decreased plasma ferritin levels in both the high-frequency and very high-frequency groups; 3) decreased plasma IgG, IgA and IgM levels in the very high-frequency and high-frequency groups and plasma albumin in the very high-frequency group only; 4) had no effect on blood pressure, body composition nor exercise performance, whatever the intensity of the donation.

This is the first randomized controlled trial prospectively investigating the health consequences of repeated plasma donation. Most research studies in this domain have an observational study design, with different study limitation (e.g. not controlled for confounding), resulting in high uncertainty on the (causal) link between repeated plasma donation and health consequences.^22–25^ One previous non-randomized controlled study compared total serum protein, albumin, IgG, IgA and IgM levels after weekly or bi-weekly (>14 days) plasmapheresis for 6 months to levels obtained in regular blood donors. ^8^ This study showed that total protein and IgG levels in the weekly group were lower than those in the control and the bi-weekly group, but remained well within the normal ranges. Albumin, IgA and IgM levels were not modified by plasmapheresis. Our high-frequency group, corresponding more or less to the weekly group in Ciszewski et al, had lower plasma ferritin, IgG, IgA and IgM levels after 3 months plasma donation. Our very high-frequency group was even more impacted, with haemoglobin and albumin levels being downregulated as well compared to the start of plasma donation and compared to the placebo group. Except for IgG in the very high-frequency group, the drop in all other haematological and biochemical parameters in the high-frequency and/or very high-frequency group did not cross the lower acceptable limits in average. Of note, before the start of the study, IgG levels were slightly lower in the low- and very-high frequency group compared to the placebo group but still well within physiological range. Lower ferritin values were previously reported in frequent plasma donors compared to non-donors, ^11 13^ with ferritin levels being negatively correlated to the number of donations per year. ^11^ When looking at individual values for ferritin levels, 5 volunteers out of 16 in the very high-frequency group had values lower than 12µg/l at the end of the 3 months. Those results contrast with a previous study retrospectively looking at ferritin levels over a period of 12 months, during which plasma was donated at frequencies ranging from 0 to more than 70 times. ^12^ Less than 1% of male donors presented ferritin levels below 12µg/l, even in the group donating at least 70 times over 12 months, approaching the frequency in our very high-frequency group. Divergent results have been reported as well concerning haemoglobin levels, with one study reporting lower levels in frequent plasma donors^11^ and another no effect^13^, even when donating at least once a week for 12 months. Finally, all studies having investigated IgG found reduced levels in donors, with frequencies from once a month to twice a week, with a higher risk at falling below the normal range. ^6 8 10 13^ Here, we found that IgG levels were reduced even in the low-frequency group, donating once a month, while IgA and IgM levels were decreased only in the high-frequency and very high-frequency groups. Globally, haematological and biochemical parameters were severely impacted by repeated plasma donation in the very high-frequency group, only mildly in the high-frequency group and not impacted in the low-frequency group compared to the placebo group.

Alongside determining the effects of repeated plasma donation on haematological and biochemical markers, the second aim of the study was to investigate the functional and physiological impact of repeated donation. We found no effect of 3-month plasma donation on blood pressure, body mass, body composition, markers for endurance performance and maximal strength. Systolic and diastolic blood pressure were found to be decreased after 4-month plasma donation at intervals of less than 14 days in donors with high baseline blood pressure levels. ^7^ Here, in donors with normal blood pressure levels at the start of the study, no modification of blood pressure was observed, even in the most intensive groups. Based on the decrease in some haematological parameters and a previous study looking at the effect of one single plasma donation on exercise performance^17^, we could have expected a decrease in markers for endurance performance in the very high-frequency group. Neither maximal power output, maximal oxygen consumption, nor blood lactate levels were modified by repeated plasma donation, whatever the intensity of the donation. One previous study looked at the effects of one single plasma donation on time to exhaustion, maximal oxygen consumption and markers for anaerobic capacity, i.e. blood lactate levels and maximal accumulated oxygen deficit. ^17^ While maximal oxygen consumption was unaffected by plasma donation, time to exhaustion, blood lactate levels and maximal accumulated oxygen deficit were all decreased by 10 to 20%. Our results suggest that the negative effects of acute plasma donation on endurance performance are not observed on the longer term, when donation is repeated and performance is determined a few days after the last donation in basal conditions. It is important to highlight that, despite the downregulation of key haematological parameters, endurance performance was not affected. We previously found that a partial dissociation and/or delay may be found between the regulation of haematological parameters, and more particularly those related to the iron status, and endurance performance after repeated blood donation. ^18 26^ Given the importance of iron status for exercise performance^27^, it might not be excluded that the 3-month period of investigation was too short to induce detectable downregulation of endurance and strength performance in the very high-frequency group.

Finally, no serious adverse events were reported and only few events (anemia, hematomas, vasovagal reactions without syncope) were present in the high-frequency and very high-frequency groups, which are classically reported by others after plasma donation. ^6 14 15^

### Clinical implications

Very high-frequency donation affects IgG levels of donors down to a level that may impact their immune system and affects haematological parameters. Therefore, countries should rely on large donor bases with donors donating at low-frequency to ensure a sustainable plasma supply while avoiding negative health effect to their donors.

### Limitations

Firstly, only middle-aged men have been included in this study. Therefore, the external validity is limited and the present results cannot be extrapolated to female or elderly donor populations. Further studies should investigate whether those results are similar in women and older people.

Secondly, this randomized controlled trial lasted for 3 months. It may not be excluded that the severe haematological and biochemical changes measured during this period in the very-high frequency group, although not falling below normal values for most of them, will not affect physiological function and exercise performance on a longer term. Thirdly, the randomization procedure was suboptimal since allocation to the high-frequency/very high-frequency group was sometimes in conflict with the availability of the participant. Therefore, the donor was assigned to the first available position on the randomization list that did not conflict with his availability. The impact of this suboptimal randomization procedure was considered to be limited.

### Conclusion

Haematological and biochemical parameters were severely impacted when plasmapheresis was repeated twice a week, mildly impacted with a frequency of three times per month and not impacted with a frequency of once a month. Despite those changes, markers for exercise performance were not altered when plasmapheresis, whatever the frequency, was repeated over 3 months.

## Data Availability

All data produced in the present study are available upon reasonable request to the authors

## ACKNOWLEDGMENT

We thank Anna Piperi, Marine Buchet and Elena De Bock for technical assistance and Céline Bugli for the statistical analyses. The study was funded by the Science Foundation of the Belgian Red Cross.

## Authorship contributions

Protocol design: VC, PV, LD; data acquisition: AM, JK, SvDdtR, CL, NB, NA, SC, VC, LD; data interpretation: HVR, PV, VC, LD; manuscript drafting: AM, LD; manuscript revision and approval: AM, JK, SvDdtR, CL, NB, NA, SC, HVR, PV, VC, LD.

## Disclosure of conflict of interest

No conflict of interest to be disclosed.

---

**Table S1.** p-values from the global ANOVA analyses.

|  | Group | Time | G*T | LF vs P | HF vs P | VHF vs P | HF vs LF | VHF vs LF | VHF vs HF |
| --- | --- | --- | --- | --- | --- | --- | --- | --- | --- |
| **RBC** | 0.7714 | 0.8316 | 0.0001 | 0.5340 | 0.6592 | 0.7527 | 0.8537 | 0.3422 | 0.4430 |
| **Haemoglobin** | 0.2233 | 0.9179 | <0.0001 | 0.4014 | 0.8031 | 0.1079 | 0.2702 | 0.4275 | 0.0606 |
| **Haematocrit** | 0.2768 | 0.4828 | 0.0006 | 0.4319 | 0.8595 | 0.1209 | 0.3298 | 0.4320 | 0.0809 |
| **Reticulocyte** | 0.0625 | 0.0032 | 0.0277 | 0.8766 | 0.0309 | 0.0771 | 0.0413 | 0.1006 | 0.6755 |
| **Iron** | 0.0023 | 0.7698 | 0.3998 | 0.3015 | 0.0573 | 0.0003 | 0.3674 | 0.0058 | 0.0560 |
| **Ferritin** | 0.0093 | 0.0003 | 0.0014 | 0.9074 | 0.0183 | 0.0101 | 0.0223 | 0.0124 | 0.8161 |
| **CRP** | 0.5313 | 0.2867 | 0.9484 | 0.2674 | 0.1925 | 0.6533 | 0.8415 | 0.4983 | 0.3807 |
| **Glycemia** | 0.2163 | 0.0322 | 0.0361 | 0.3968 | 0.8885 | 0.2266 | 0.4736 | 0.0396 | 0.1728 |
| **Insulinemia** | 0.7541 | 0.0155 | 0.4047 | 0.9930 | 0.5582 | 0.3752 | 0.5585 | 0.3732 | 0.7594 |
| **HbA1c** | 0.7651 | 0.0962 | 0.0007 | 0.5003 | 0.5150 | 0.8803 | 0.9813 | 0.4026 | 0.4157 |
| **CK** | 0.4041 | 0.4766 | 0.4489 | 0.7384 | 0.9659 | 0.1424 | 0.7684 | 0.2498 | 0.1497 |
| **Total cholesterol** | 0.3562 | <0.0001 | 0.3901 | 0.9402 | 0.7967 | 0.1395 | 0.7355 | 0.1153 | 0.2131 |
| **Albumin** | 0.0193 | 0.0002 | <0.0001 | 0.2828 | 0.5907 | 0.0035 | 0.5835 | 0.0520 | 0.0139 |
| **IgA** | 0.5834 | <0.0001 | <0.0001 | 0.1852 | 0.4879 | 0.7155 | 0.5162 | 0.3259 | 0.7370 |
| **IgG** | <0.0001 | <0.0001 | <0.0001 | 0.0044 | 0.0091 | <0.0001 | 0.7874 | 0.0029 | 0.0013 |
| **IgM** | 0.6411 | <0.0001 | <0.0001 | 0.9522 | 0.5045 | 0.2738 | 0.5368 | 0.2928 | 0.6616 |
| **SBP** | 0.5625 | 0.0615 | 0.1499 | 0.4532 | 0.2608 | 0.5819 | 0.2608 | 0.1888 | 0.8481 |
| **DBP** | 0.7827 | 0.4888 | 0.9039 | 0.3464 | 0.8636 | 0.5864 | 0.4344 | 0.6830 | 0.7060 |
| **Body mass** | 0.5687 | 0.4193 | 0.3678 | 0.6449 | 0.1703 | 0.4591 | 0.3509 | 0.7755 | 0.5158 |
| **BMI** | 0.8068 | 0.2363 | 0.3311 | 0.8858 | 0.3662 | 0.7542 | 0.4391 | 0.8632 | 0.5469 |
| **BMC** | 0.8843 | 0.3905 | 0.0803 | 0.5429 | 0.6718 | 0.9546 | 0.8505 | 0.4990 | 0.6253 |
| **Fat mass** | 0.3698 | 0.0002 | 0.7008 | 0.0819 | 0.6340 | 0.3163 | 0.7617 | 0.9787 | 0.9954 |
| **FFM + BMC** | 0.5218 | 0.0119 | 0.7919 | 0.2558 | 0.1870 | 0.6342 | 0.8495 | 0.4990 | 0.8495 |
| **FFM** | 0.5051 | 0.0124 | 0.7596 | 0.2492 | 0.1751 | 0.6160 | 0.8328 | 0.5050 | 0.3810 |
| **%Fat** | 0.2492 | 0.0002 | 0.7500 | 0.4295 | 0.3613 | 0.3625 | 0.0864 | 0.0868 | 0.9980 |
| **Quadriceps 1RM** | 0.8626 | 0.3709 | 0.4308 | 0.9542 | 0.7622 | 0.4464 | 0.8035 | 0.4748 | 0.6405 |
| **Arm max strength** | 0.5259 | 0.4520 | 0.5672 | 0.6111 | 0.3487 | 0.9299 | 0.1445 | 0.5446 | 0.3875 |
| **Lactate 190W** | 0.6069 | 0.8933 | 0.9188 | 0.5094 | 0.6642 | 0.5529 | 0.8125 | 0.2127 | 0.3016 |
| **Lactate post** | 0.1126 | 0.0618 | 0.8807 | 0.1553 | 0.3513 | 0.4126 | 0.6130 | 0.0250 | 0.0781 |
| **Max power output** | 0.6435 | 0.0055 | 0.5343 | 0.7727 | 0.7668 | 0.3566 | 0.5524 | 0.2203 | 0.5237 |
| **VO<sub>2</sub>peak** | 0.2584 | 0.0014 | 0.7932 | 0.3942 | 0.1223 | 0.0668 | 0.4745 | 0.3100 | 0.7621 |
| **VE<sub>max</sub>** | 0.8337 | 0.7687 | 0.8645 | 0.5660 | 0.8345 | 0.3992 | 0.7103 | 0.7839 | 0.5186 |
| **HR<sub>max</sub>** | 0.3332 | 0.7756 | 0.9383 | 0.2922 | 0.9427 | 0.4481 | 0.3182 | 0.0684 | 0.3991 |
| **Oxygen pulse** | 0.2920 | 0.0015 | 0.5445 | 0.9715 | 0.8069 | 0.1093 | 0.7761 | 0.0969 | 0.1661 |

RBC, red blood cell; CRP, C-reactive protein; HbA1c, glycated haemoglobin; CK, creatine kinase; IgA, immunoglobulin A; IgG, immunoglobulin G, IgM, immunoglobulin M; SBP, systolic blood pressure; DBP, diastolic blood pressure; BMI, body mass index; BMC, bone mineral content; FFM, fat free mass; 1RM, 1-repetition maximum; VO<sub>2</sub>peak, peak oxygen consumption; VE, ventilation; HR, heart rate; RER, respiratory exchange ratio; P, placebo; LF, low-frequency; HF, high-frequency; US, US scheme. n=15 in Placebo, n=16 in Low-frequency (LF), High-frequency (HF) and Very high-frequency (US).

**Table S2.** Subjects characteristics at the start of the study.

|  | Placebo |  | Donation |  | p-value |
| --- | --- | --- | --- | --- | --- |
|  |  | Low-frequency | High-frequency | Very high-frequency |  |
| **Age (y)** | 35.9±2.0 | 37.7±2.2 | 31.4±1.8 | 31.6±2.1 | 0.071 |
| **Body mass (kg)** | 77.4±3.8 | 78.9±2.6 | 82.8±2.1 | 80.7±2.9 | 0.595 |
| **Height (cm)** | 180.7±2.0 | 182.6±2.0 | 184.4±1.9 | 183.0±2.0 | 0.720 |
| **BMI (kg·m<sup>-2</sup>)** | 23.6±0.9 | 23.7±0.6 | 24.4±0.7 | 24.0±0.6 | 0.840 |
| **Sport/week (h)** | 5.4±0.8 | 5.9±0.9 | 5.0±0.7 | 4.9±0.9 | 0.620 |
| **VO<sub>2</sub>peak (ml·kg<sup>-1</sup>·min<sup>-1</sup>)** | 49.0±2.8 | 46.3±2.3 | 44.3±1.3 | 44.1±1.9 | 0.364 |
| **Donation history** | 0[2] | 0[1] | 1[2] | 2[4] | 0.281 |

Values are means ± SEM, except for donation history where the number of plasma donations until 3 months prior to the study was expressed as a median [IQR, interquartile range]. A mixed model was used for all variables to test differences between groups except for donation history, for which a Kruskal-Wallis test was used. BMI, body mass index; VO<sub>2</sub>peak, peak oxygen consumption. n=15 in Placebo, n=16 in Low-frequency, High-frequency and Very high-frequency.

**Table S3.**
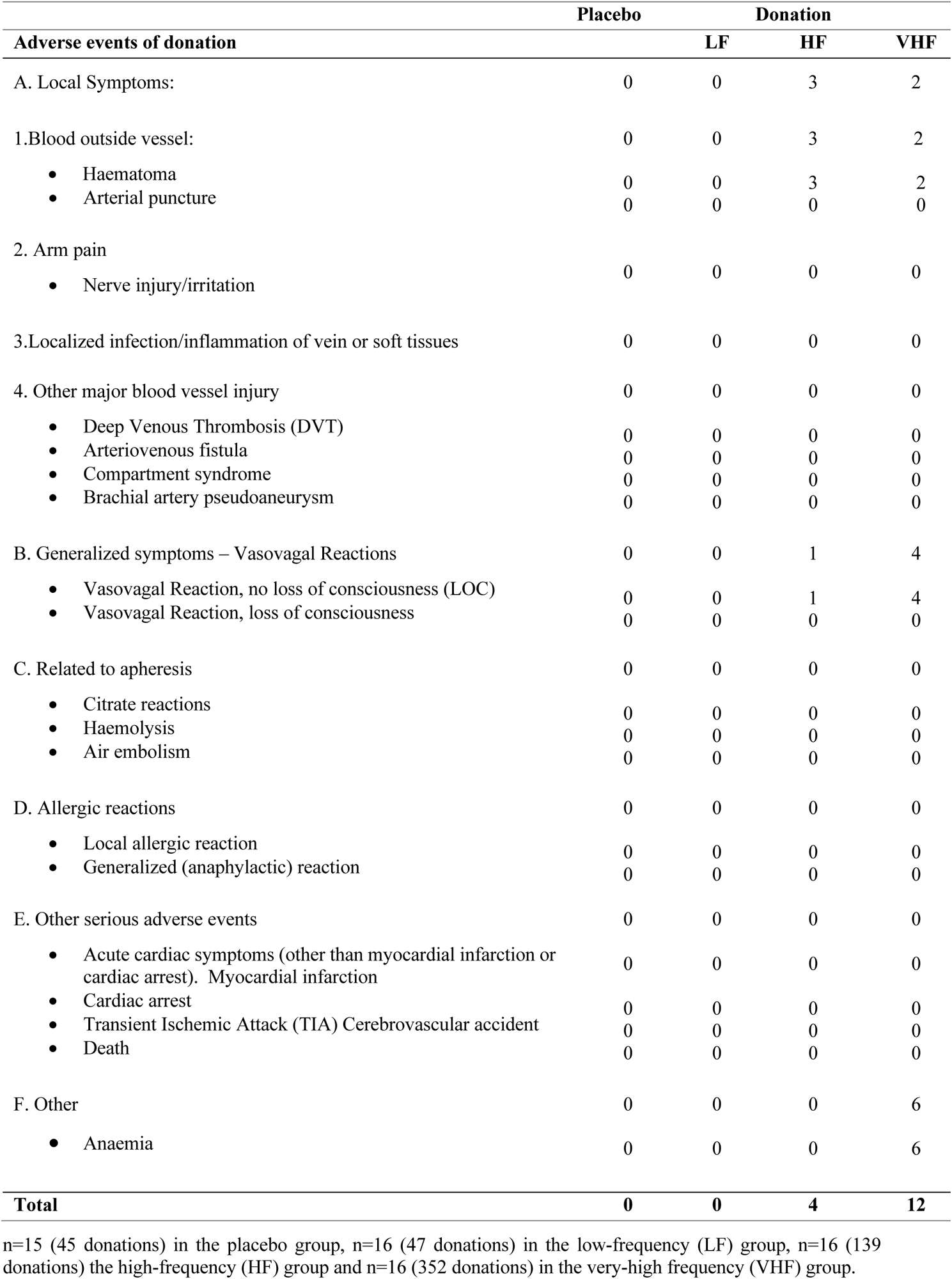
Number of clinical adverse events in each group.

## Notes

### Competing Interest Statement

The authors have declared no competing interest.

### Clinical Trial

NCT05815615

### Clinical Protocols

https://register.clinicaltrials.gov/prs/app/action/SelectProtocol?sid=S000D0C3&selectaction=Edit&uid=U00069FK&ts=2&cx=-br52hk

### Author Declarations

The study was approved by the Ethics Committee of UCLouvain and the investigation was performed according to the principles outlined in the Declaration of Helsinki.

